# Efficacy of a spatial repellent for control of malaria in Indonesia: a cluster-randomized controlled trial

**DOI:** 10.1101/19003426

**Authors:** Din Syafruddin, Puji BS Asih, Ismail Ekoprayitno Rozi, Dendi Hadi Permana, Anggi Puspa Nur Hidayati, Lepa Syahrani, Siti Zubaidah, Dian Sidik, Michael J. Bangs, Claus Bøgh, Fang Liu, Evercita C. Eugenio, Jared Hendrickson, Timothy Burton, J. Kevin Baird, Frank Collins, John P. Grieco, Neil F. Lobo, Nicole L. Achee

## Abstract

A cluster randomized, double-blinded, placebo-controlled trial was conducted to estimate protective efficacy of a spatial repellent against malaria infection at Sumba, Indonesia. Following radical cure in 1,341 children aged ≥ 6 months - ≤5 years in 24 clusters, households were given transfluthrin or placebo passive emanators (devices designed to release vaporized chemical). Monthly blood screening and biweekly human-landing mosquito catches were performed during 10-months baseline (June 2015 to March 2016) and a 24-month intervention period (April 2016 to April 2018). Screening detected 164 first-time infections and an accumulative total of 459 infections in 667 subjects in placebo-control households; and 134 first-time and 253 accumulative total infections among 665 subjects in active intervention households. The 24-cluster protective effect of 27.7% and 31.3%, for time to first-event and overall (total new) infections, respectively, was not statistically significant. Purportedly, this was due in part to zero to low incidence in some clusters, undermining the ability to detect a protective effect. Subgroup analysis of 19 clusters where at least one infection occurred during baseline showed 33.3% (*p*-value = 0.083) and 40.9% (*p*-value = 0.0236, statistically significant at the 1-sided 5% significance level) protective effect to first-infection and overall infections, respectively. Among 12 moderate-to high-risk clusters, a statistically significant decrease on infection by intervention was detected (60% protective efficacy). Primary entomological analysis of impact was inconclusive. While this study suggests spatial repellents prevent malaria, additional evidence is required to demonstrate the product class provides an operationally feasible and effective means of reducing malaria transmission.

## INTRODUCTION

It has been nearly 75 years since the role of spatial repellency (deterrency or avoidance) was first described as a potentially beneficial attribute in malaria control, showing chemicals could effectively disrupt normal host-seeking mosquito behavior and interrupt contact with humans, thus preventing disease transmission.^1-3^ Spatial repellency is used here as a general term to refer to a range of insect behaviors induced by airborne, volatile chemicals that ultimately result in a reduction in human-vector contact. These behaviors include movement away from a treated space with chemical stimulus, interference with host detection (attraction-inhibition) and/or interference with feeding response (feeding-inhibition).^4-7^ Spatial repellency can be measured and distinguished from other chemical actions, primarily contact irritancy and toxicity, in the laboratory^8, 9^ and in semi-field evaluations.^10, 11^ Which behavior is elicited by a volatile chemical depends on the concentration or dose-exposure interaction by the mosquito in the treated space. For example, toxicity occurs at higher chemical doses while deterrence (behavioral avoidance) can result from lower, sub-lethal chemical concentrations.^12^ Currently, the majority of commercial spatial repellent products utilize either low concentrations of short-duration United States Environmental Protection Agency (USEPA) registered synthetic pyrethroids (pyrethrin, metofluthrin and more recently transfluthrin)^13^ or botanical-based compounds.^14, 15^

The World Health Organization (WHO) Vector Control Advisory Group (VCAG) assesses evidence on the epidemiological effectiveness of new vector control interventions and by doing so supports WHO’s development of policy recommendations, including the potential use of spatial repellents as a public health vector control strategy.^16^ Reviewed by VCAG since 2014, the assessment of rigorous epidemiological evidence for endorsing a policy recommendation of a spatial repellent intervention class remains limited and deemed insufficient.^17^ A malaria prevalence study in China evaluating mosquito coils containing 0.03% transfluthrin demonstrated a 77% reduction in human *Plasmodium falciparum* cases^18^ and the use of coils containing 0.00975% metofluthrin provided 52% protective efficacy (PE) against new (incident) malaria infections in Indonesia.^19^ Although findings from both studies were encouraging, neither met the required VCAG evidence for full pubic health assessment either due to lack of adequate scale in study design (small cluster number in Indonesia) and/or being underpowered (both studies).^20^ The importance of a WHO policy for implementation of spatial repellents in malaria control programs could dramatically increase investments by private industry to develop chemicals that operate through modes of actions to elicit vector behavior changes other than purely toxicity. This would potentially introduce a new generation of effective active ingredients and product formulations into the disease control/eradication arsenal.^21, 22^ In combination with existing WHO-recommended malaria control interventions, spatial repellents may add protective benefit in reducing vector-borne disease.^23^ This is most highlighted in settings where traditional long-lasting insecticidal nets (LLINs) or indoor residual spraying (IRS) may not be sufficiently protective due to varying circumstances: 1) early-evening blood feeding vectors; ^24, 25^ 2) when LLINs are not in use or used intermittingly; ^26, 27^ and/or 3) where vectors do not or limit resting time indoors on insecticide-treated surfaces;^24, 25^ 4) be unavailable, or are impractical and/or infeasible such as during humanitarian emergency relief operations.^28^ Control or elimination of malaria in these circumstances will require innovative approaches; as example spatial repellents providing a highly beneficial protective role against transmission.^29, 30^

The current claim of a spatial repellent intervention class for public health value is *‘deployment of a spatial repellent will prevent human-vector contact to reduce pathogen transmission’*. The role of spatial repellency in public health is dependent on assessing whether or not human protection is rigorously evidenced, thus the requirement of clinical trials using a prototype spatial repellent product for the intervention class. With this epidemiological primary endpoint in mind, the objectives of the current randomized cluster trial (RCT) were to build upon previous epidemiological findings and provide rigorous evidence of a spatial repellent that provides a sufficient protective effect against malaria infection risk and demonstrate risk reduction by decreasing relevant entomological measures (e.g., anopheline human landing (biting) density, age-grading parity and/or sporozoite infectivity rates) in endemic communities. The rationale for conducting the RCT at Sumba Island stems from an essential requirement by WHO VCAG that more evidence of human health impact is needed to recommend a WHO global public health spatial repellent policy^31^ and that such evidence should come from varied malaria endemicity settings (low, moderate and high).^16^ Sumba Island was selected as the study site based on the following criteria: 1) active malaria transmission, 2) vector populations exhibiting early-evening and/or outdoor biting patterns, and 3) housing characteristics which reflect probability of indoor exposure to mosquitoes. The underlying reason for integrating entomological measures (above) in the RCT was to identify correlations, if any, with PE which could then be used in future trials with appropriate minimum thresholds required to demonstrate non-inferiority of next-generation spatial repellent products.

## METHODS

The study was conducted from June 2015 to April 2018, registered in clinicaltrials.gov (Identifier: NCT02294188) and performed according to The International Conference on Harmonisation’s (ICH) Guideline for Good Clinical Practice (GCP; document E6) (R1) and The OECD Principles of Good Laboratory Practice (GLP) based on The National Agency of Drug and Food Control, Indonesia regulations.^32, 33^ There were no changes to methods after trial commencement.

### Ethics statement

Ethical review and approval for this study was granted by the Ethics Committee (EC) of the Faculty of Medicine, Universitas Hasanuddin (Protocol #UH14070385), the University of Notre Dame (Protocol #14-01-1448) and endorsed by the Eijkman Institute Research Ethics Committee, Jakarta, Indonesia. Consent was obtained from parents or guardians of child recruits following EC guidelines. For sentinel households participating in entomological collections, signed consent was sought from the head-of-household. No data on individuals was collected during the entomological collections. During the consenting process, the study was described and the relevant consent form read in local dialect by study staff. The consent form detailed the design of the study including radical cure, the purpose of collecting and storage of blood samples (towards laboratory-based malaria diagnosis), descriptions of the study risks, benefits, and procedures of therapeutic radical cure and follow-up. For households recruited for entomology measures, the consent form detailed the design of mosquito collections and descriptions of study risks, benefits and procedures of human landing catch (HLC).

Participants who were illiterate were asked to appose their thumbprint and, when possible, a literate witness was asked to sign (i.e., a witness selected/known by the participant and having no connection to the research team). All households were provided with a signed copy of the consent form after agreeing to participate in the study. Adverse events were captured during participant follow-up and entomological collections and reported to monitoring authorities in accordance with the approved protocol. All participants were free to drop out of the study at any time, regardless of reason or having to provide one, and without prejudice.

### Study setting

The study was conducted in Southwest and West Sumba districts, East Nusa Tenggara Province, Indonesia (**Figure 1**). The circa 400,000 residents of the two districts occupy 175 village ‘groups’ (*desas*) and several small-sized towns.^34^ Thirteen village groups, with resident populations ranging between 1,067 and 3,904 (avg. 2,132), served as study locations for the final selection of the 24 study clusters consisting of multiple desas. Organized bed net campaigns were recently introduced with the last mass distribution occurring in February-March 2018 across the study area. A mass distribution of long-lasting insecticidal nets (LLINs) occurred in October-December 2014 in both districts (Olyset Net® (Olyset Net®, permethrin 2.0% w/w) with a reported > 95% coverage of households provided 1-4 nets each. A second round of LLIN mass distribution occurred February-March 2018 (PermaNet® 3.0, deltamethrin 180mg/m2 + PBO synergist). The last round of focal IRS occurred in 2003 using a pyrethroid-based product. The malaria prevalence based on microscopically diagnosed parasitemia drawn from a random sample of 50% of residents present in 13 villages conducted in 2015, 10 months before the intervention trial, averaged 15.5% (2.5% to 37.3%) (**Table 1**).

**Table 1.**
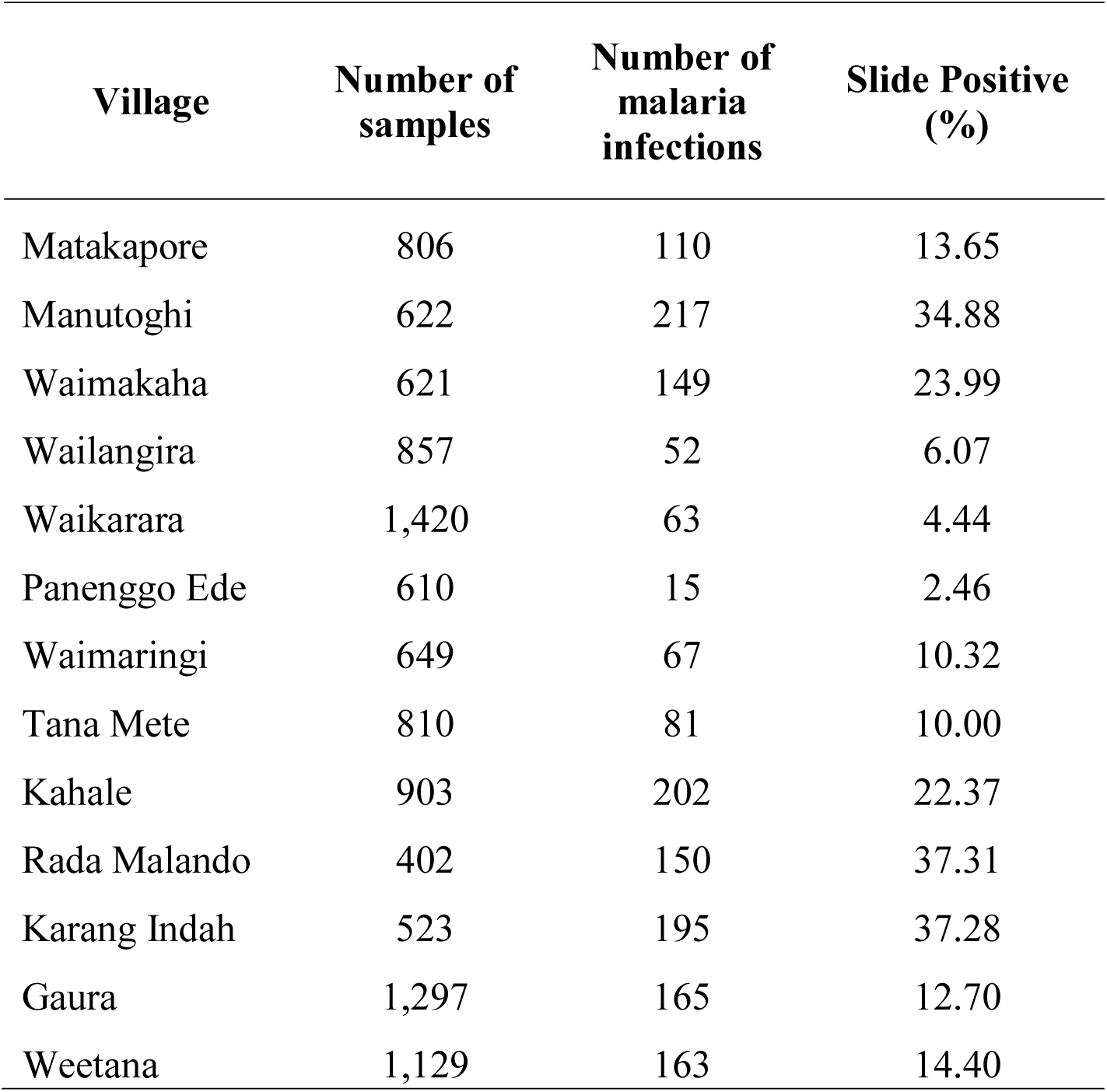
Malaria infection prevalence data from 10 months before randomization in the 13 villages from which study clusters were delineated on Sumba Island.

| **Village** | **Number of samples** | **Number of malaria infections** | **Slide Positive (%)** |
| --- | --- | --- | --- |
| Matakapore | 806 | 110 | 13.65 |
| Manutoghi | 622 | 217 | 34.88 |
| Waimakaha | 621 | 149 | 23.99 |
| Wailangira | 857 | 52 | 6.07 |
| Waikarara | 1,420 | 63 | 4.44 |
| Panenggo Ede | 610 | 15 | 2.46 |
| Waimaringi | 649 | 67 | 10.32 |
| Tana Mete | 810 | 81 | 10.00 |
| Kahale | 903 | 202 | 22.37 |
| Rada Malando | 402 | 150 | 37.31 |
| Karang Indah | 523 | 195 | 37.28 |
| Gaura | 1,297 | 165 | 12.70 |
| Weetana | 1,129 | 163 | 14.40 |

**Figure 1.**
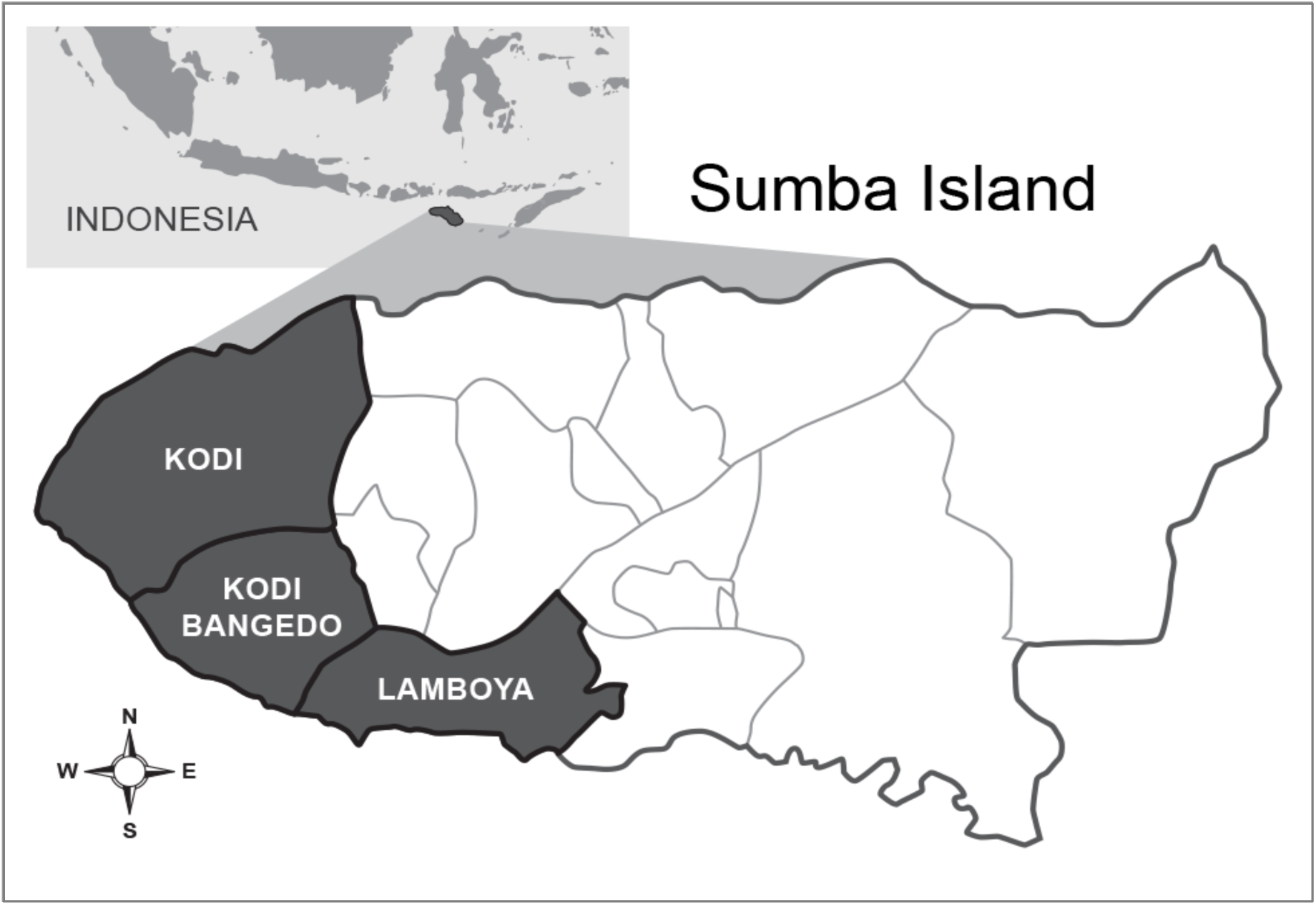
Study site areas in Southwest and West Sumba districts located in Kodi, Kodi Bangedo and Lamboya sub-districts of Sumba Island, Nusa Tenggara Timur Province (eastern Lesser Sunda islands), Indonesia (map not to scale).

Although very little detail is known about the malaria vector distribution and bionomics in this study area,^35^ an earlier (August 2007) entomologic survey documented 13 species of anophelines occurring in West Sumba district: *Anopheles aconitus* (Dönitz), *Anopheles annularis* Van der Wulp, *Anopheles barbirostris* s.l. (Satoto), *Anopheles flavirostris* (Ludlow), *Anopheles hyrcanus* group (Reid), *Anopheles indefinitus* (Ludlow), *Anopheles kochi* (Dönitz), *Anopheles leucosphyrus* group (Reid), *Anopheles maculatus* Theobald, *Anopheles subpictus* s.l. Grassi, *Anopheles sundaicus* s.l. (Dusfour et al.), *Anopheles tessellatus* Theobald, and *Anopheles vagus* Dönitz.^36^ These species vary spatially in relative abundance associated with presence of their preferred aquatic larval habitats ranging from coastal brackish and freshwater marshes and ponds, seasonally productive rice paddies, to forested hillsides with perennial running streams and small rivers. At the time of the survey, human-landing collections revealed *An. subpictus* and *An. vagus* as the predominant species in the upland interior locations, and *An. sundaicus* as the most common species attracted to human along the coastal plain. The majority of residents in study villages work as small-holder agriculturalist pursuits, typically lacking public supplied electricity or articulated water supply systems. Homes are predominately traditional designed, large, thatched-roof structures raised ∼1m above ground, averaging ∼70 m^3^ in size (6 m length × 6 m width × 2 m in height) and constructed with gaping bamboo walls and flooring that offer little protection from mosquito entry (**Figure 2**).

**Figure 2.**
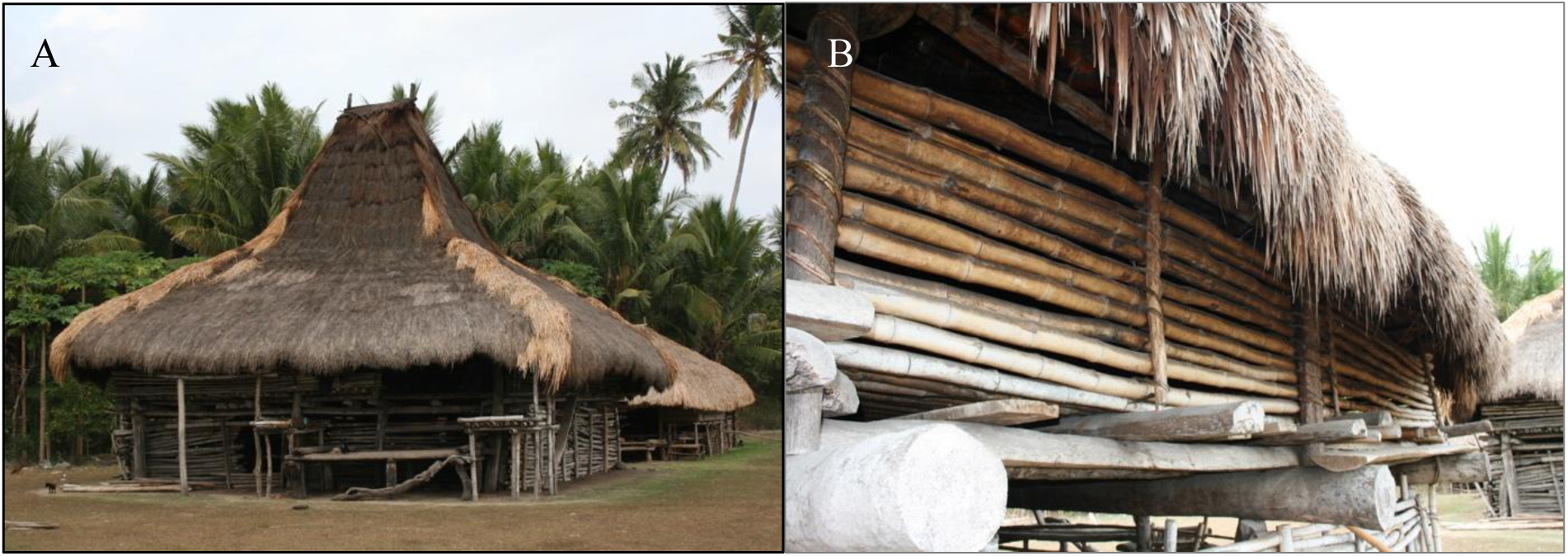
Traditional Sumba house structure (A) raised ∼1m above ground and averaging ∼70 m^3^ in size with thatch roof, bamboo floors and walls (B), which offer minimal protection from mosquito entry.

### Trial design

The study was a randomized cluster, double-blind, placebo-controlled, clinical trial with a total of 24 clusters divided into 12 clusters per intervention arm. Clusters were selected based on housing type, focusing on traditional houses (with or without thatch roofing) (**Figure 2**), mitigation of potential chemical dispersion ‘spillover’ effect (i.e., distance between nearest homes in different clusters ∼500 m apart), and logistical considerations (i.e., distance from field-based satellite laboratories). About fifty-four households were recruited within each cluster based on human sample size requirements. Households from 13 villages were stratified into 24 clusters before randomization: Gaura (pop. 2,831; houses 432), Kahale (pop. 3,904, houses 253), Karang Indah (pop. 1,169, houses 139), Manutoghi (pop. 1,067; houses 165), Matakapore (pop. 2,805; houses 209), Panenggo Ede (pop. 1,271; houses 90), Rada Malando (pop. 1,842; houses 133), Tana Mete (pop. 2,124; houses 213), Waikarara (pop. 2,709; houses 446), Wailangira (pop. 1,878; houses 226), Waimakaha (pop. 2,334; houses 211), Waimaringi (pop. 2,598; houses 116), and Weetana (pop. 2,670; houses 275) (**Figure 3**).

**Figure 3.**
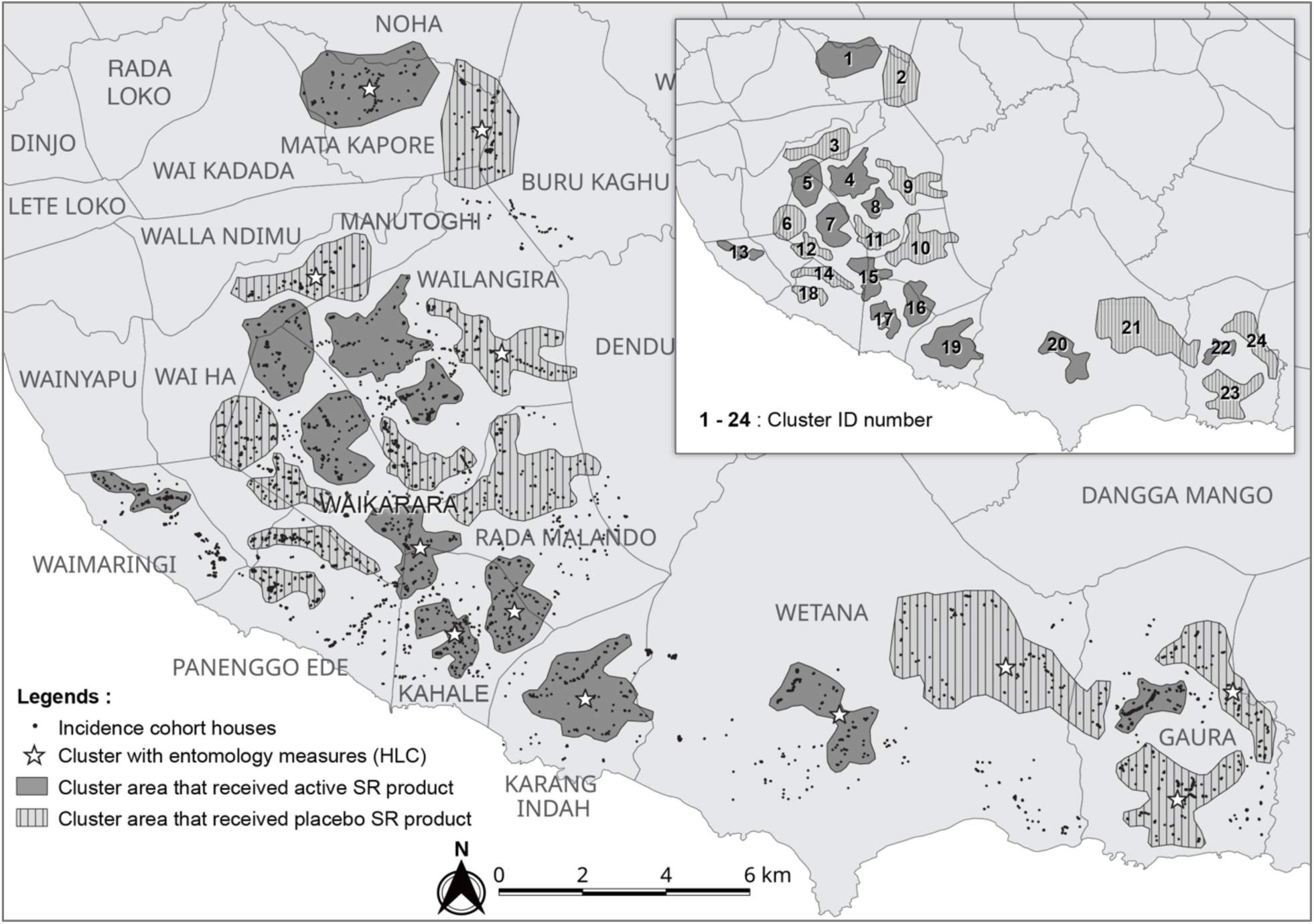
Location of 24 study clusters in West and Southwest districts, Sumba. Clusters were selected for enrolling the incidence cohort, each consisted of ca. 100 households with an average distance of 500m between clusters. A total of 48 sentinel houses from 12 clusters were selected for routine entomological human landing catch (HLC).

#### Sample size

Previous malaria incidence rates collected in a portion of the study area in coastal Kodi Subdistrict were used to estimate the likely malaria attack rate in the current study villages at 0.3 infections/person-year.^19^ Sample size determination was based on the hazard rate comparison in the proportional hazards regression model.^37^ The required number of first-time infections was estimated at 417 to permit detection of a 30% protective effect by the spatial repellent intervention compared to placebo with 80% power at the type-I error rate, assuming a between-cluster coefficient of variation (CV; defined as the ratio between standard deviation and mean) of 30% and a baseline hazard rate of 0.3 per person-year. With 12 clusters per treatment arm (active or placebo), with 2-month accrual and 22 months follow-up, and an estimated 20% loss-to-follow-up (LTFU) during intervention, a total of 54 subjects per cluster was required (n=1,296). As entomology measures were used in relationship analyses of PE rather than hypothesis testing, sample size was based on field logistical capacity and not for statistical power.

### Participants

The average number of children ≤5 years of age in each cluster was 68 (57-79). From individual households in study clusters, one child, aged ≥6 months - ≤5 years at time of recruitment, were provided the opportunity to enroll as subjects in the study. Following informed consent, medical screening consisted of physical examination by a study physician and a qualitative NADPH spot test for G6PD deficiency (Trinity Biotech qualitative G6PD assayTM, ref 345-UV, Trinity Biotech, St. Louis, MO).^38^ Additional to being G6PD normal status, eligibility requirements included a bodyweight ≥ 40 kg, hemoglobin > 5mg/dL (Hb201+, HemoCue AB, Angelholm, Sweden),^39^ no severe acute illness/infection on the day of inclusion, temperature ≤ 38°C, participant acknowledging sleeping in the village > 90% of nights during any given month, not participating in another clinical trial, and with no intensions for extended travel during the study period.

### Intervention

A 24-month intervention follow-up period was implemented from April 2016 to April 2018. Intervention was simultaneously initiated in all subject households and non-subject households that consented to receive intervention. The spatial repellent intervention is a transfluthrin-based passive emanator produced by S.C. Johnson & Son, Inc (SCJ) designed to release a volatile chemical into the air without the requirement of an external heat source, such as electricity or fire, and prevent human–vector contact by disrupting normal behavior of mosquitoes at a distance within the treated space. The indoor placement of the intervention product was designed with the objective to measure PE under indoor use conditions.

Transfluthrin is a registered compound commonly found in commercially available mosquito coils globally based on WHO specifications.^40^ The U.S. Environmental Protection Agency (EPA) has recently approved transfluthrin products for indoor use within the USA.^41^ Emanators (active and placebo) of identical packaging and color were distributed by study personnel every 2 weeks at the individual household application rate of 2 units / 9m^2^ according to SCJ specifications. There was a median of 1 room (range 1,9) and 10 emanators (range 4,56) per household. Intervention devices were positioned indoors by hanging individual emanators on two metal hooks specially attached to walls for this study. Each position remained static throughout the study. The hooks facilitated stabilization of the interventions so the chemical-treated surface was consistently exposed facing the interior space. Research staff placed, removed and replaced emanators in households at set intervals and recorded attrition, based on number of products removed from each structure during each replacement period, to ascertain application rate for use in estimating cluster coverage. During the trial period, quality control analysis was performed by Ross Laboratories, India on unused (in storage) emanators to verify amount of transfluthrin in actives and the absence of transfluthrin in placebos. At the end of follow-up period (April 2018), used, unused and expired emanators were disposed of by PT. Wahana Pamunah Limba Industri in Jakarta according to Indonesian regulations on disposal requirements.

### Randomization, allocation and blinding

Clusters were allocated to receive either active or placebo treatment using a random number generator (https://www.random.org). The cluster allocation code was made available from the intervention manufacturer to the Data Safety Monitoring Board (DSMB) for use in safety assessments. The site database manager assigned a unique identification number to each household (HIN) and the site intervention administrator coordinated distribution of blinded active or placebo to enrolled households within each cluster corresponding to the pre-labeled package code. Unblinded assignments were shared with a site administrator in a sealed envelope placed in a secure location within the managing center of the research project (Jakarta) for purposes of emergency unblinding related to adverse and serious adverse events. Thus, the investigators, research team, study subjects, and residents were blinded as to which cluster received active versus placebo devices until after completion of the study.

### Procedures

#### Radical cure and follow-up

**Figure 4** summarizes screening, enrollment, and follow-up of the incidence density cohorts. The trial consisted of a 10-months baseline follow-up period (June 2015 to March 2016) and a 24-month intervention follow-up period (April 2016 to April 2018). A total of 1,353 subjects were presumptively radically treated using a fixed combination formulation of dihydroartemisinin (DHA)-piperaquine (P) (containing 40 mg dihydroartemisinin and 320 mg piperaquine (Zhejiang Holley Nanhu, Beijing Holley Cotec) administered as a weight per dose regimen of 2.25 and 18 mg/kg per dose of dihydroartemisinin and piperaquine, respectively, once daily for 3 days. Primaquine (PT, Pharos Tbk, Semarang, Indonesia) 0.25 mg/kg body weight was prescribed for the 14 days immediately before implementing intervention. The DHA-P combination is currently the first-line antimalarial drug for uncomplicated malaria treatment in Indonesia.

**Figure 4.**
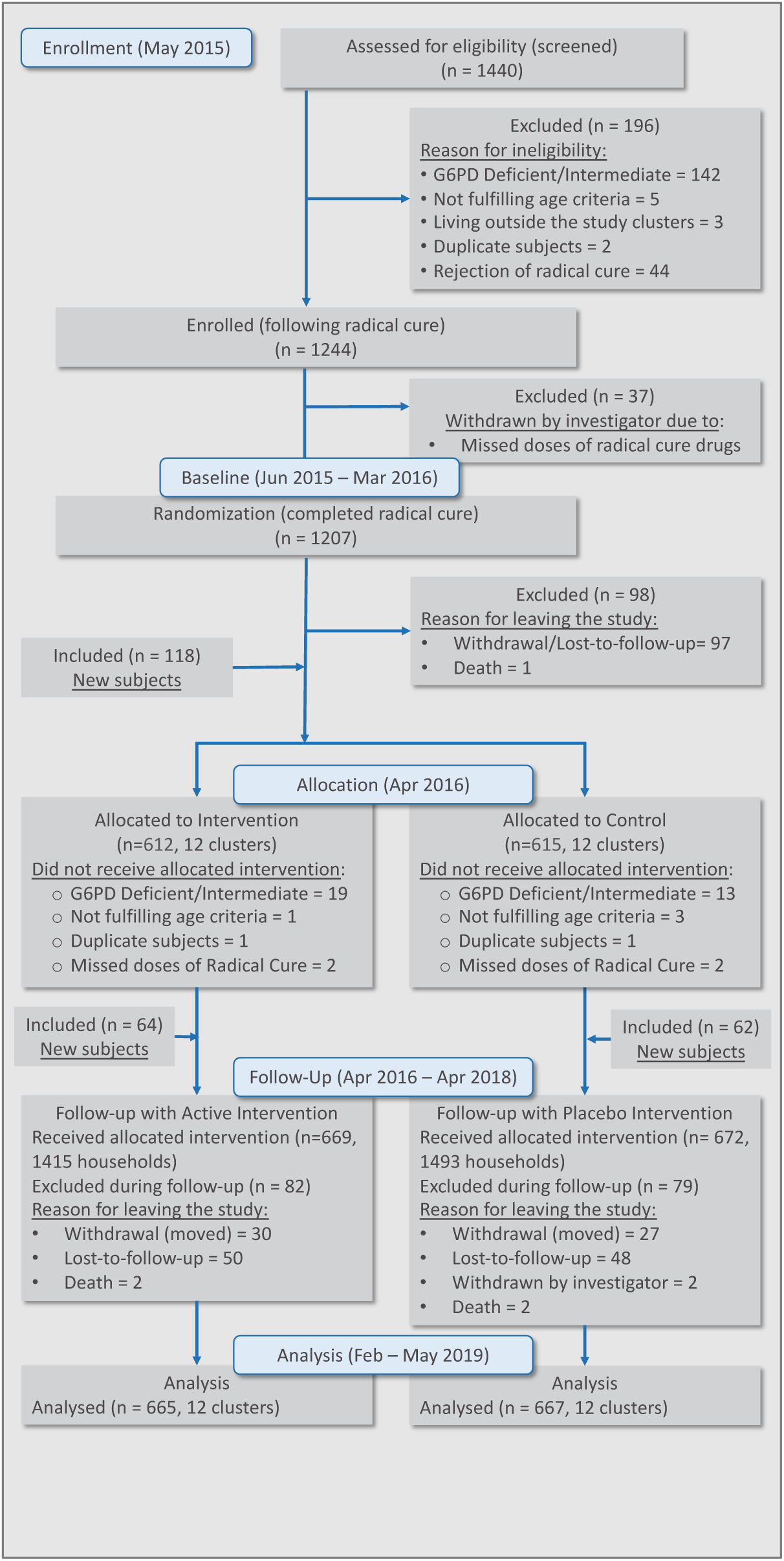
Flowchart of enrollment of study volunteers.

All subjects were examined for *Plasmodium* spp. infection by expert microscopy, and later confirmed using polymerase chain reaction (PCR) from matched filter paper blood spot samples.^42^ New malaria infections among the participants were monitored every 4 weeks using microscopic examination of Giemsa-stained blood films according to WHO guidelines based on a minimum of 200 high magnification (1000x oil-immersion) thick blood film fields.^43^ Two certified expert microscopists independently (blinded) examined slides on-site at project-dedicated field laboratories. PCR detection of parasite DNA was conducted at the Eijkman Institute for Molecular Biology (EIMB) central laboratory in Jakarta for all four *Plasmodium* species.^42^ A blood sample was defined as malaria ‘positive’ for inclusion in incidence analyses if meeting criteria of having two diagnostic outcomes indicating presence of parasite (e.g., 2x microscopy; 1x microscopy + 1x PCR; or 2x PCR). All positive and 10% randomly selected negative samples diagnosed at EIMB were re-tested at the University of Notre Dame. Discordant microscopy and PCR results required reexamination of both initial findings. Participants found malaria infected at point-of-care were immediately treated with DHA-P and were removed from contributing further to person-time at risk of first-time infection; however, they remained in the study to monitor overall (total new) incidence for malaria infections and the expected number of infection cases averted.

#### Entomologic surveys

In a subset of 12 clusters, adult mosquito diversity and densities were measured using human-landing catches (HLC) every two weeks from start of baseline through end of follow-up period in a subset of 12 clusters. Clusters for entomological sampling were selected based on exhibiting highest HLC densities during baseline sampling, along with existence of mosquito larval habitats. The 12 clusters were hierarchically stratified on criteria of baseline human landing catch then blindly allocated to treatment arm to ensure a balanced recruitment (6 clusters in each treatment group). Four neighboring sentinel houses within each of the 12 clusters were randomly selected for mosquito collections (n=48). Collections were conducted at sentinel houses for 1 night every 2 weeks from paired active/placebo clusters (e.g., 3 pairs on Monday night and 3 pairs on Wednesday night) during intervention. Teams of two collectors were assigned per house, one positioned indoors near the center of the house and one located outside on the house verandah, approximately 1 m from the exterior wall. Collectors removed all mosquitoes landing on their exposed lower legs using a mouth aspirator. Collections were conducted from 1800 to 0600 h for 50 min every hour. Paired collectors rotated between indoor and outdoor positions each hour. Samples were placed into individual holding containers labeled by collection hour, unique house code (linked to blinded treatment code), and collection location (indoor or outside). Captured mosquitoes were immediately killed by ether-soaked cotton pads in the field and initially identified to species (or species complex) using morphological characters.^44^ All specimens were transported to an on-site base laboratory upon completion of the 12 h collection and a random sample of representative anopheline species (up to 20% per cluster per indoor/outdoor location and hour of collection) were dissected for parity and scored as either gravid/parous or nulliparous.^45^ Partial (head-thorax for those dissected for parity) or whole anopheline specimens were placed singly into individual Eppendorf® 1.5ml vials and stored over silica gel desiccant until further processing at EIMB, Jakarta, for detection of malaria sporozoites and molecular-based species identification, where applicable.^46, 47^ Mosquito samples were evaluated for *Plasmodium* species infection using polymerase chain reaction (PCR) methodologies to derive corresponding malaria sporozoite infection rates by parasite and vector species.^48^ Together with time-adjusted HLC densities (anophelines/person-night), matched sporozoite rates were used to derive the entomological inoculation rates (EIRs) for each treatment arm.^49^ Anopheline species identification was verified at the University of Notre Dame following previous protocols (not reported herein).^46, 47^

#### Insecticide susceptibility assays

Permethrin was evaluated using the WHO standard tube and CDC bottle assay during baseline, intervention and post-intervention periods at the WHO recommended discriminating concentration for anophelines 0.75% and CDC recommended dose 21.5µg/ml active ingredient.^40, 41^ Both WHO tube assays and CDC bottle assays were performed on F0 mixed anopheline species collected as immatures in 13 of the 24 study clusters from across 23 habitat locations during baseline, intervention and 6-mo post-intervention period (ending Oct 2018). Assays were conducted using non-bloodfed, 3-5 day old females according to established guidelines.^50, 51^ After each test period, all chemical and control specimens were stored individually over silica gel for analysis at EIMB to confirm species identification and for detection of target-site mechanisms (e.g., *kdr* gene mutations) of resistance (not reported herein).

#### Monitoring of adverse events (AE) and serious adverse events (SAE)

AEs, possibly related to transfluthrin exposure, in subjects and other household members, were captured by the study team during both active and passive blood sampling using a standardized survey form.

Investigation of study-related AEs was performed by the on-site study clinician. SAEs were also recorded according to protocol, regardless of possible relationship to intervention. Government clinic health records were compiled on a quarterly basis starting in December 2017 for DSMB safety assessment of the study population.

#### Data management and verification

Data collection was designed around a tablet-based survey platform linked to a custom-built database and web portal. CommCare (Dimagi Inc, MA, USA) was selected as the frontend form application, providing critical capabilities, including: a parent-child case structure, the ability to store forms when offline, update form versions after deployment, build forms with complex logic in a web browser, and export form data to other tools. Data was cleaned according to rules specified in the study protocol to ensure data integrity. Study data related to participating subjects and households, intervention (placement/replacement activity), mosquitoes collected, and lab analyses was cross-checked, identifying missing, incomplete, or suspect data submissions. These data were relayed to the site data manager to be resubmitted or corrected. Once data correction was complete, data was verified and requested for analyses.

### Outcomes and statistical methods (S1. Statistical Analyses Plan)

The *primary analysis* of the study was intent-to-treat, and included all the recruited subjects per their treatment assignment. Study participants were excluded from the analyses either when they had no blood samples during the intervention period or when a household might have contributed more than two subjects. In the latter case, this occurred (rarely) when there was loss-to-follow-up (LTFU) on the first recruited subject and a second subject was recruited as a replacement; only the subject with a longer follow-up period was used based on per-protocol allowing only one subject per household.

The main objective of the study was to demonstrate and quantify the protective efficacy (PE) of a spatial repellent product in reducing the incidence of malaria infection in a human cohort. Since the time to malaria in this study was measured and interval-censored, we compared the hazard rate, which is the instantaneous incidence rate, of the first-time infection and the overall infections between spatial repellent and placebo to address the study epidemiological objectives. The hazard rate is the main, and often the only, effect measure reported in many epidemiologic studies, including studies on malaria. ^52-55^

The *primary hypothesis* on PE against first-time malaria infection was tested by comparing the hazard rates of the first-time malaria infection between active and placebo (blank) intervention. The complementary log-log (cloglog) regression model log 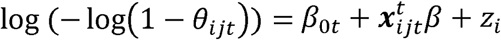 was.^56-59^ *θ*_*ijt*_ is the discrete time hazard rate of subject j in cluster *I* at time *t*, x_*ijt*_ contains visit (as a categorical predictor), the individual-level (age, gender), household-level (number of doors, open eaves Y or N, wall type), and cluster-level (baseline incidence rate, cluster population size, intervention group) covariates, and z_i_ ∼ N(0, σ^-2^) is the cluster-level random effect. PE was estimated by 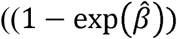 100% with a 90% confidence interval (CI) based on the Wald test, where 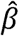 is the estimated regression coefficient associated with the intervention group, and exp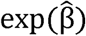is the estimated hazard ratio (HR) between active and placebo. The null hypothesis that PE is 0 was tested by the Wald’s test 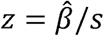 where s is the estimated standard error of 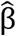 It was concluded that active intervention reduces the first-time malaria hazard rate compared to placebo if z < z_O.O5_ = -1.645; otherwise, the study would not have sufficient evidence to suggest that active intervention reduces the first-time malaria hazard rate compared to placebo at a one-sided significance level of 5%. The Kaplan-Meier (KM) curves on time to first infection per cluster was provided for the active and placebo arms, respectively.

The statistical analysis of the overall (total new) malaria infections detected in study subjects was similar to that used for analyzing the primary endpoint of the first-time malaria infection except that the above cloglog model has additional term z_j(i)_, which is the random effect at the individual level to account for the dependence among multiple malaria infections per individual. The same set of analyses as described above were performed in a subgroup analysis in clusters with non-zero baseline incidence rates and those clusters with entomology data.

The effects of active intervention on the *secondary entomological endpoints* were estimated using a negative binomial regression model, if applicable. Specifically, the *anopheline landing rate* (surrogate ‘bite’ based on HLC and indicator of human-vector contact) is defined as the number of mosquitos captured during a 12-hr evening interval (1800-0600). The covariates in the model include the fixed effects of intervention group, the interaction between treatment and location of collection (inside or outside), visit (as categorical), baseline incidence rate, baseline vector count, cluster population, and random effects for household nested within cluster and for cluster. The percentage change in human landing rate by intervention was estimated from the model. The frequencies and percentages of captured anophelines were also summarized by species. The set of covariates in the model for analyzing the *parity rate* are similar to the landing rate model, with an additional offset term for the daily landing rate. Due to data sparsity for the *sporozoite positivity rate* (> 99% of tested mosquitos were uninfected), no model-based analysis was performed and only summary statistics are provided. The *entomological inoculation rate (EIR)* is defined as the number of malaria infective mosquito bites a person receives per a unit time (typically annually) and calculated as EIR = sporozoite positivity rate × human biting rate. The observed data sparsity experienced with low sporozoite positivity also impacts the EIR results, and thus only summary statistics are provided for EIR.

Besides the model-based analyses on the hazard rate of malaria infection, we also estimated the first-time incidence rate and the overall-time infection rate over the 2-year study period. The first-time incidence rate is calculated as the number of first-time infections divided by the person-time at risk for the first-time infection (the time taken to the first malaria infection summed across all subjects in the study). The overall-time infection rate was calculated as the number of total malaria infections divided by the person-time at risk for malaria infection (the time taken to any new malaria infection, summed across all subjects during the whole study). The difference in the overall (total new) incidence rate per person-year between spatial repellent and placebo can be regarded as the total number cases averted per person-year.

## RESULTS

### Protective effect against malaria infection (pre-planned)

Trial outcomes show baseline covariates regarding subject, house construction, population and baseline malaria incidence between the active and placebo arms were balanced at the individual-, household-, and cluster-level in the all 24 cluster analyses (**Table 2**). The intervention coverage rate, defined as the proportion of actually placed emanators over the total number required per household, ranged from 82.2% to 98.6% by cluster over the entire intervention period, with a mean application rate of 93.2% and 92.3% for the active and placebo arms, respectively. The percent of LLIN usage per household during the trial, defined as responding ‘Yes’ to the question: *“Did you use a bed net last night?”*, ranged from 14.6% to 99.8% in clusters that received active intervention and 17.4% to 99.8% in clusters that received placebo (**S2. LLIN Usage**).

**Table 2.** Summary baseline covariates for both spatial repellent (SR) and placebo treatment arms for the primary analysis.

| **Individual level** |  |  |
| --- | --- | --- |
|  | **SR (n = 665)** | **Placebo (n = 667)** |
| Age in months (mean ± SD, (min, max)) | 34.0 ± 15.1 (6, 59) | 34.2 ± 14.8 (6, 59) |
| Gender (% male subjects) | 54.1% | 51.1% |
| **Household level** |  |  |
|  | **SR (n = 665)** | **Placebo (n = 667)** |
| House wall type (wood %) | 91.6% | 94.0% |
| Open eaves (yes %) | 98.0% | 99.6% |
| No. of doors (mean ± SD, (min, max)) | 2.06 ± 0.38 (1, 4) | 2.06 ± 0.30 (0, 4) |
| **Cluster level** |  |  |
|  | **SR (n = 12)** | **Placebo (n = 12)** |
| Cluster population (mean ± SD, (min, max)) | 694.3 ± 59.2<br>(624, 820) | 730.8 ± 129.0<br>(616, 1117) |
| Baseline incidence rate per person-year (mean ± SD, (min, max)) | 0.096 ± 0.115<br>(0, 0.426) | 0.089 ± 0.088<br>(0, 0.265) |
| Baseline overall (total new) infection incidence per person-year (mean ± SD, (min, max)) | 0.094 ± 0.111<br>(0, 0.412) | 0.089 ± 0.087<br>(0, 0.261) |

There were 134 first-time infections among 665 subjects in active intervention households and 164 first-time infections in 667 subjects in placebo-control households, with a 27.7% decrease in the first-time malaria hazard risk using active compared with placebo (90% CI: -21.3%, 56.9%) (**Table 3**). The 27.7% PE was not statistically significant at the 5% one-sided significance level (*p* = 0.151). The estimated PE of active intervention against overall (total) malaria infections (first and subsequent) was 31.3% (90% CI: -10.8, 57.4%) with a one-sided *p*-value = 0.098 (**Table 3**). Investigation of potential shift in parasite species infection frequency (*P. falciparum* vs. *P. vivax*) between active and placebo arms will be reported in subsequent publications.

**Table 3.** Summary first-time and overall (total new) malaria incidence during study.

|  | First-time infection |  |  | Overall (total new) infections |  |  |
| --- | --- | --- | --- | --- | --- | --- |
|  | All clusters | Subgroup 1<sup>#</sup> | Subgroup 2<sup>*</sup> | All clusters | Subgroup 1<sup>#</sup> | Subgroup 2<sup>*</sup> |
|  | **SR : Placebo** |  |  |  |  |  |
| No. of clusters | 12:12 | 10:9 | 6:6 | 12:12 | 10:9 | 6:6 |
| No. of households | 665 : 667 | 557 : 500 | 335 : 335 | 665 : 667 | 557 : 500 | 335 : 335 |
| No. of infections | 134 : 164 | 130 : 158 | 93 : 140 | 253 : 439 | 249 : 433 | 196 : 408 |
| Person years | 1079 : 1032 | 873 : 722 | 506 : 444 | 1216 : 1216 | 1015 : 909 | 609 : 607 |
| Incidence rate (per person-year) | 0.124 : 0.159 | 0.149 : 0.219 | 0.184 : 0.315 | 0.208 : 0.361 | 0.245 : 0.476 | 0.322 : 0.672 |
| Hazard Ratio (90% CI) | 0.723<br>(0.431, 1.213) | 0.64<br>(0.299, 1.367) | 0.447<br>(0.302, 0.664) | 0.687<br>(0.426, 1.108) | 0.591<br>(0.382, 0.914) | 0.344<br>(0.233, 0.508) |
| PE (%) (90% CI) | 27.7<br>(-21.3, 56.9) | 33.3<br>(-7.8, 58.7) | 55.3<br>(33.6, 69.9) | 31.3<br>(-10.8, 57.4) | 40.9<br>(8.61, 61.8) | 65.6<br>(49.2, 76.7) |
| 1-sided *p*-value | 0.151 | 0.083 | 0.0004 | 0.098 | 0.0236 | <0.0001 |
| <sup>#</sup> Subgroup 1: clusters with non-zero incidence during intervention |  |  |  |  |  |  |
| <sup>*</sup> Subgroup 2: clusters with entomology data collected (moderate-to-high risk of infection) |  |  |  |  |  |  |

A total of 164 first-time malaria infections occurred during approximately 1032 person-years at risk in study participants whose households were given placebo, with a calculated incidence density of 0.159 infections/person-year (**Table 3, Figure 5**).^60^ In contrast, 134 total malaria attacks occurred in transfluthrin-active households with approximately 1079 person-years at risk resulting in a calculated 0.124 infections/person-year. A cumulative 439 malaria infections (first-time and subsequent) during approximately 1216 person-years at risk in participants with households provided placebo, produced an incidence density of 0.361 infections/person-year. Contrastingly, 253 total (accumulative) malaria attacks occurred among participants living in active intervention households with approximately 1216 person-years at risk equaling 0.209 infections/person-year (**Table 3**).

**Figure 5:**
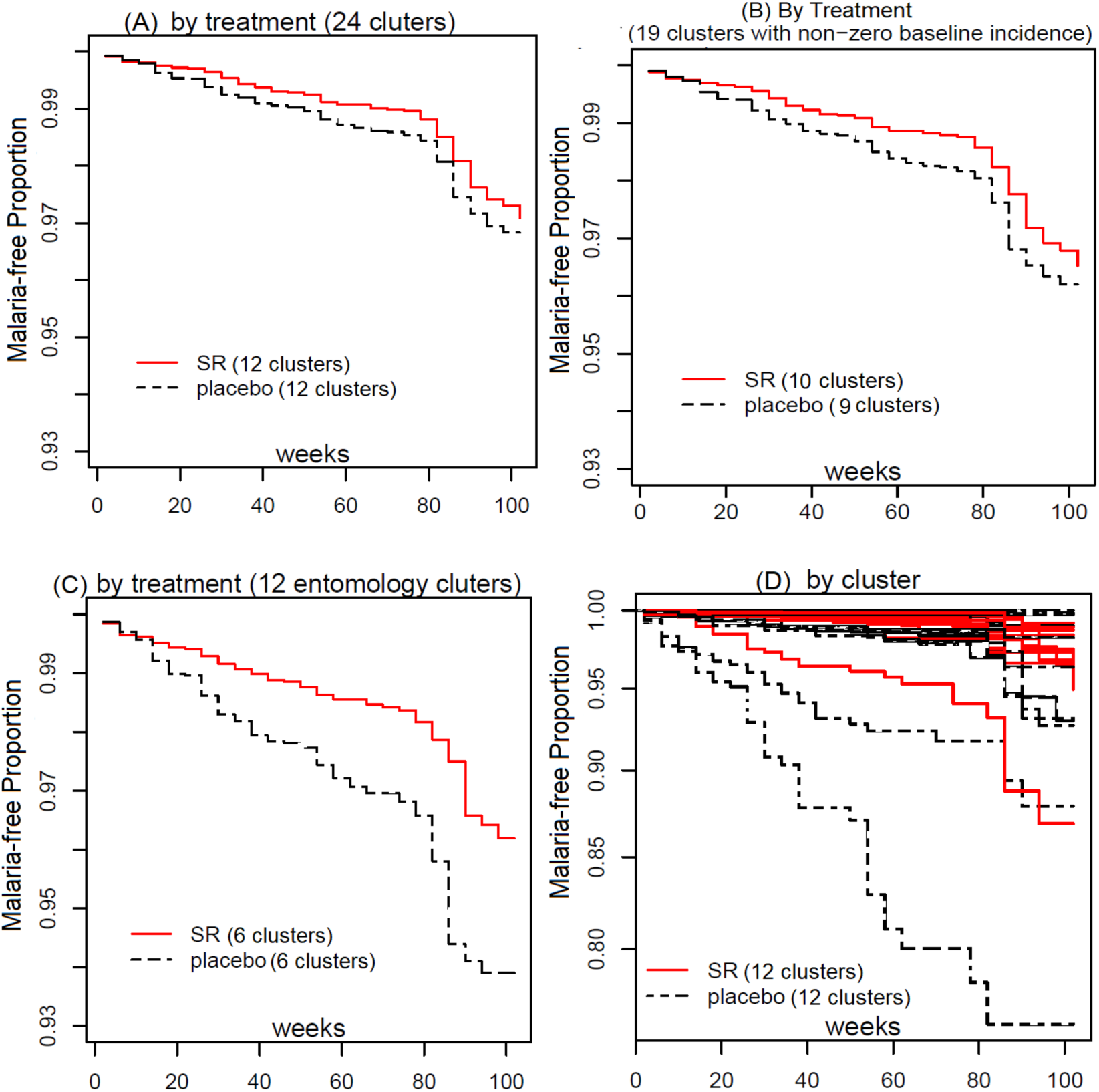
Kaplan-Meier curves for (A) treatment (spatial repellent intervention and placebo treatment arm) for each subset analyses (B and C) and for all study clusters (D).

### Subgroup analyses of protective effect against malaria infection among non-zero incidence and entomology clusters (not pre-planned)

Among the 24 clusters, there were five clusters with zero baseline incidence having had zero to very low incidence rates during intervention. The first subgroup analysis used the 19 clusters with non-zero incidence rate as the remaining five clusters may have been excluded from the intervention if baseline analysis had been performed before randomization. Excluding these five clusters, the estimated PE for overall (total new) infections in the remaining subgroup of 19 clusters with non-zero baseline incidence rates was 40.9% (90% CI: 8.61, 61.8%) resulting in a one-sided *p*-value of 0.0236 (**Table 3**). The second subgroup analyses included incidence from the 12 clusters having routine entomology collection data (i.e., mean anopheline landing rate/person), and where the average baseline incidence was approximately 4 folds greater than the other study clusters. The PE using active intervention against time to first-event and overall (total new) malaria infections in this subgroup was 55.3% (90% CI: 33.6, 69.9; *p*-value <0.0004) and 66% (90% CI: 49.2%, 76.7%; *p*-value <0.0001), respectively (**Table 3**). The data indicate the baseline covariates (subject, house construction, population and baseline malaria incidence) between the active and placebo arms were balanced at the individual-, household-, and cluster-level in the 19 non-zero incidence and 12 entomology cluster subgroup analyses (not shown).

### Effects on entomological endpoints (pre-planned)

Results presented are summary outcomes on aggregated anopheline species captured using HLC. Detailed data analyses on effects of this intervention on entomology measures is forthcoming in subsequent publications, to include temporal change in species-specific anopheline vector composition over the trial period, relative abundance between treatment arms by species, and the HLC of non-anophelines (culicine mosquitoes).

#### Anopheline landing rates

A total of 52 weeks of HLC were performed within a subset of 12 clusters during the intervention period. Results based on morphological species identification detected 19 putative malaria vector species showing spatial and temporal variation across monitored clusters. The most common anopheline species (s.s. or s.l.) attracted to humans included: *Anopheles aconitus, An. annularis, An. barbirostris, An. flavirostris, An. kochi, An. maculatus, An. subpictus, An. sundaicus, An. tessellatus* and *An. vagus* (**S3. Anopheline Frequency Summary**). The cumulative indoor (n=8,780) and outdoor (n=9,207) anopheline landing rates across both baseline and intervention sampling periods are shown in **Figure 6**. There was a numerical reduction in indoor and outdoor landing density from collections performed in households provided active intervention (n= 3,883 and 4,373, respectively) compared with placebo households (n= 4,897 and 4,834, respectively). The reduction in anopheline attack rate on collectors positioned indoors and outdoors, at sentinel households with active intervention compared with placebo houses was not statistically significant: 16.4% (95% CI = -75.2%, 182.7%; *p* = 0.774) and 11.3% (95% CI = -73.7%, 199.4%; *p* =0.847), respectively (**Table 4**).

**Table 4:** Intervention effect on anopheline mosquito landing rates by collection location.

| **Location<sup>1</sup>** | **SR**<br>**(mean ± SD)<sup>2</sup>** | **Placebo**<br>**(mean ± SD)** | **% Change (95% CI)**<br>**SR vs. Placebo** | **2-sided**<br>**p-value** |
| --- | --- | --- | --- | --- |
| Indoor | 3.14 ± 5.84 | 3.97 ± 8.73 | -16.4 (-75.2, 182.7) | 0.774 |
| Outdoor | 3.54 ± 7.13 | 3.93 ± 8.64 | -11.3 (-73.7, 199.4) | 0.847 |
| <sup>1</sup> Position at each human landing catch (HLC) sentinel house where sample was captured (indoor = near center of house; outdoor = on verandah ~1m from edge of exterior wall). |  |  |  |  |
| <sup>2</sup> Human landing rate based on 12-hour collection per person. |  |  |  |  |

**Figure 6.**
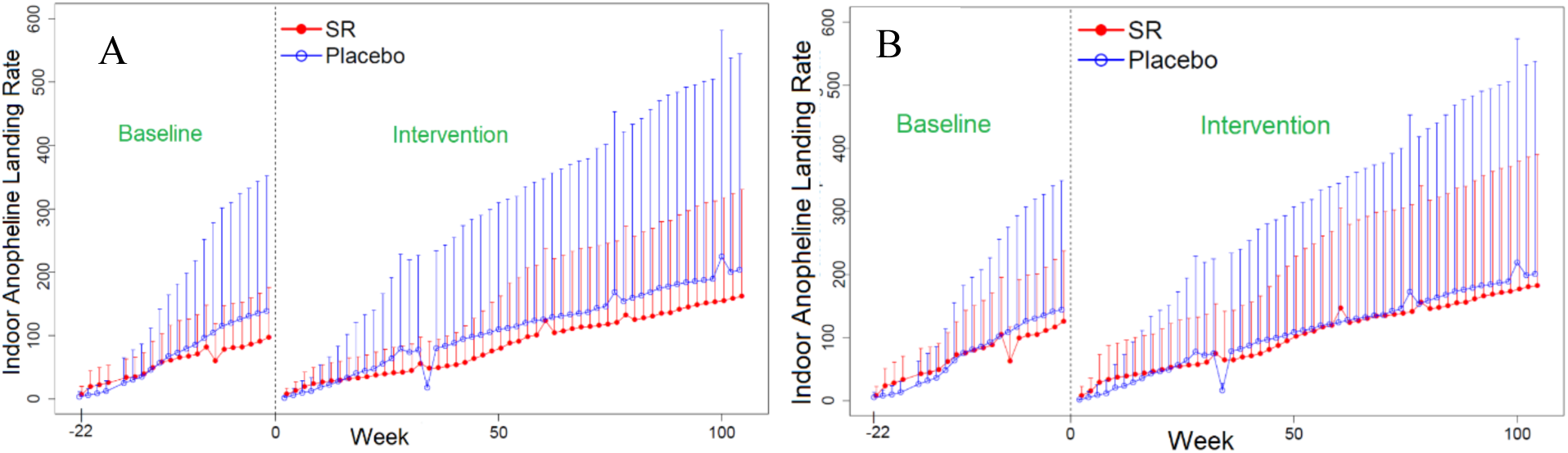
Mean (+ SD) cumulative biweekly indoor (A) and outdoor (B) anopheline human landing catch (HLC) averaged over 20 to 24 households per treatment arm - spatial repellent (SR) intervention and placebo, respectively.

#### Sporozoite positivity rate and Entomological Inoculation Rate (EIR)

A total of 11,928 and 17,986 anopheline samples from baseline and intervention follow-up periods, respectively, were processed for malaria sporozoites. The frequency of sporozoite positive anophelines is provided in **Table 5**. Only *P. falciparum* and *P. vivax* infections were detected in captured mosquitoes. During baseline and intervention period, the sporozoite rate was less than 0.5% for both treatment and placebo arms. Data sparsity regarding comparison of sporozoite rates precluded inferential statistical analyses. The EIR was <1 bite per year in both treatment arms during baseline and intervention periods.

**Table 5.** Frequency of anopheline (all species) sporozoite infection status for both spatial repellent (SR) and placebo treatment arms.

| **Treatment Allocation** | **Pf<sup>1</sup>** | **Pv<sup>2</sup>** | **Indeterminate<sup>3</sup>** | **Non-infected** | **Sporozoite Positivity Rate<sup>4</sup>** |
| --- | --- | --- | --- | --- | --- |
| **Baseline** |  |  |  |  |  |
| SR | 12 | 9 | 0 | 4706 | 0.44% |
| Placebo | 12 | 9 | 0 | 6244 | 0.34% |
| **Intervention** |  |  |  |  |  |
| SR | 3 | 8 | 0 | 8130 | 0.14% |
| Placebo | 6 | 1 | 1 | 9615 | 0.07% |
| <sup>1</sup>Pf = *Plasmodium falciparum*<br><sup>2</sup>Pv= *Plasmodium vivax*<br><sup>3</sup>Indeterminate defined as *Plasmodium* positive sample but parasite species not identifiable<br><sup>4</sup>Sporozoite positivity rate = [(Pf + Pv) / (Pf + Pv + Indeterminate + non-infected)] x 100% |  |  |  |  |  |

#### Parity rate (age-grading)

A total of 15,828 anopheline samples were dissected for parity status during the trial period (**Table 6**). The proportion of females categorized as “older,” combining parous and gravid states (mosquitoes with advanced ovarian follicle development as evidence of a recent blood meal), and those “younger” as nulliparous (non-bloodfed) and more recently emerged, were compared between active and placebo treatments for the 12 clusters with entomological monitoring was conducted. Overall, transfluthrin-active emanators proportionally increased nulliparity in the sampled anopheline populations compared to placebo for both indoor and outdoor locations (**Table 6**).

**Table 6.** Spatial repellent (SR) intervention effect on parity and nulliparity rates (all anopheline species).

|  | **HLC Location<sup>1</sup>** | **SR (mean ± SD)<sup>2</sup>** | **Placebo (mean ± SD)** | **% Change (95% CI) SR vs. Placebo** | **2-sided p-value** |
| --- | --- | --- | --- | --- | --- |
| Parous | Indoor (n = 5,784) | 0.41 ± 0.44 (n = 2,537) | 0.41 ± 0.45 (n = 3,247) | -10.2 (-62.1, 113.2) | 0.808 |
|  | Outdoor (n = 6,193) | 0.40 ± 0.44 (n = 2,907) | 0.43 ± 0.45 (n = 3,286) | -25.9 (-68.8, 75.6) | 0.495 |
| Nulliparous | Indoor (n = 1,873) | 0.16 ± 0.29 (n = 994) | 0.12 ± 0.26 (n = 879) | 58.3 (-37.0, 298.0) | 0.329 |
|  | Outdoor (n = 1,978) | 0.17 ± 0.30 (n = 1,126) | 0.11 ± 0.25 (n = 852) | 54.9 (-37.6, 284.3) | 0.346 |
| <sup>1</sup> Position of collector conducting human landing catch (HLC) (indoor = near center of house; outdoor = on house verandah ~1m from edge of exterior wall).<br><sup>2</sup> Mean and SD are descriptive statistics; and % change was obtained from fitting negative binomial models. |  |  |  |  |  |

#### Insecticide susceptibility

A total of 5,091 adult female anophelines (chemical and control bioassays) were evaluated for susceptibility: 700 samples during baseline, 1,805 during intervention and 2,586 post-intervention. *Anopheles vagus* was the most widely distributed and tested anopheline (56%), followed by *An. sundaicus* (12.6%) and *An. subpictus, An. barbirostris, An. kochi, An. aconitus, An. maculatus*, and *An. tessellatus* (proportionally, all < 10%). In the CDC bottle bioassay, baseline, intervention and post-intervention tests using permethrin (21.5ng/ml) showed 100% knockdown between 15-30 min exposure and within the recommended diagnostic time. In the WHO tube bioassay with rare exception, there was 100% 24hr mortality to permethrin 0.75% following 60 min exposure. Based on the assumption of predominately pyrethroid-susceptible wild populations of *Anopheles* species present in trial sites, there was no conclusive evidence of the development of phenotypic resistance to pyrethroid class chemicals between pre-and post-intervention periods. Interestingly, in several clusters during the post-intervention phase, there were indications of reduced permethrin susceptibility (WHO bioassay) in a few *An. barbirostris* populations to permethrin. Follow-up investigations provided inconsistent results, thus confirmation that these focal populations showed low levels of resistance could not be verified. The determination of presence or absence of target-site mechanisms for resistance will be reported separately.

### Adverse Events (AE) and Severe Adverse Events (SAE)

Using government clinic health records, a total of 523 AEs were reported from non-participants while 144 AEs were reported from the intervention study cohort captured by the study team. Of subject AEs, 52 were reported from participants in the active intervention arm and 92 from placebo clusters. General respiratory complaints were most common for all reported AEs, followed by general fever. There were 6 total SAEs reported in the overall study cohort during follow-up: one death due to suspected brain infection; one death from respiratory infection; one death from malaria with concomitant bacterial or viral infection; one death by drowning, and two deaths of unknown cause.

### Intervention Quality Control

A total of 180 unused emanator samples were analyzed in 2017. Samples were taken from 12 different active and placebo clusters with manufacturing dates ranging from April to November 2016 (15mo to 8mo-old samples). All sampled placebo interventions (n = 48) were found to be absent for transfluthrin. The average transfluthrin quantity from all sampled active emanators (n= 132) was 54.3 mg, which was within the specification range (55.0 mg ± 2.75 mg).

## DISCUSSION

Malaria remains a significant global public health burden despite recent progress in reducing transmission rates.^61^ The primary objective of this large-scale RCT conducted on Sumba Island, Indonesia was to demonstrate and quantify the protective efficacy (PE) of a passive emanating spatial repellent intervention (transfluthrin-treated), for reducing malaria incidence (transmission) in humans. Such epidemiological evidence of health impact is a fundamental requirement in the critical path of development of new vector control intervention classes being assessed by the WHO VCAG.^16^ Sumba Island, Indonesia represents a group of malaria endemic settings where spatial repellents are intended to be implemented if/when a policy recommendation is endorsed as Sumba does not currently conduct routine IRS, LLINs were only recently distributed and there is a range of anopheline biting habits by local vectors, to include early-evening and/or outdoor biting.

The primary per-protocol analyses provide an estimate of 27% PE against malaria infection, which is near the targeted efficacy of 30% used in the power calculations. This effect, however, was not statistically significantly protective at the 1-sided 5% significance level. There were 298 first-time infections, in contrast to an expected number of at least 417 assumed in the sample size calculations, thus resulting on the trial being underpowered. The demonstration of spatial repellency PE (66%) against total new infections in the subset of 12 clusters used for entomological measures, which had approximately 4 folds greater malaria infection incidence during intervention, indicates health impact where risk of malaria infection was greatest (i.e., locations with highest vector landing densities). Outcomes reported here are promising for malaria control, despite the study being unable to demonstrate clear, statistically protective efficacy on primary analysis, with data anticipated to be included in meta-analyses with other clinical trials evaluating spatial repellents.

This study highlights three primary challenges for consideration in future spatial repellent trials as well for new vector control intervention classes more broadly: 1) firstly, having ‘adaptive’ study designs, especially for evaluation of interventions in low to moderate malaria transmission settings and/or settings with inherently large cluster-to-cluster variance on transmission intensity; 2) defining and identification of the ‘key’ entomological correlates of protection; and 3) ensuring reliability and feasibility in AE/SAE reporting for accurate safety assessment. Regarding adaptive study designs, our assumptions for power and sample size calculations were based on a previous proof-of-concept study that took place in an entirely coastal location but was spatially very near (bordering) the current study area.^19^ The study villages, both coastal and further inland, were selected with the assumption that all clusters delineated from these villages would experience similar levels of malaria transmission during the intervention period; however, this was not the case during the trial. As in all epidemiological trials, underlying assumptions of incidence used for power and sample size calculations can vary from actual incidence during intervention follow-up. Baseline data analyses indicated zero-incidence for five of the 24 selected study clusters with a 5-folds higher between-cluster variability than assumed in the original sample size calculation. The five clusters with zero baseline incidence (two allocated to receive active and three allocated to receive placebo) also had either zero or low incidence rates during intervention and ranked in the top 8 among the 24 clusters in terms of lowest in intervention incidence (data not shown).

In retrospect, a pre-planned interim and final analysis of the baseline data would have allowed for consideration in adjustment of the study design by filtering out the zero-incidence clusters, or increasing the study duration (time of follow-up), the number of subjects recruited per cluster, or the number of clusters to better satisfy requirements of statistical power and capture the necessary number of outcome events (infections). However, baseline incidence was measured only for verifying hierarchical stratification and use as a covariate in statistical analyses of PE. This therefore, resulted in: 1) the original sample size not accommodating for interim and final analyses of baseline data; 2) timelines of grant period not being projected and assured to include lag-time for baseline data analyses and outcome assessment; and perhaps most importantly 3) baseline incidence analyses not being explored early to determine variations in sample size /cluster number adjustments required to achieve sufficient study power and subsequent study site viability.

Given the updated baseline incidence rate of 0.131 per person-year and 97.1% CV, to maintain 80% power with the originally assumed 30% PE, 100 clusters per arm with 144 households (HHs; i.e. subjects) would have been required to be recruited to collect 5,550 first-time malaria events – an unmanageable scale that could not have been supported due to geographical, logistical and funding constraints. Moreover, increasing the follow-up time to collect more first-time malaria events would have neither counterbalanced the large variability nor the longer duration required to collect the additional events due to the seasonally-influenced low to moderate malaria transmission among clusters. For this reason, when planning future trials in low or moderate endemic settings, investigators should consider including a greater number of clusters per arm and/or building into the statistical analyses plan an interim and final analyses of *baseline* incidence with pre-determined study design adaptations, to include down-selection of clusters with predetermined incidence thresholds. This may become most relevant when considering proof of efficacy of an intervention in low transmission settings as a component for malaria elimination goals. Early exercises of the impact of a lower than assumed incidence (or greater CV) during study planning should be explored amongst investigators, industry and funding partners to ensure that the study area context, intervention manufacturing, program period and/or funding can sufficiently meet the demands of power requirements if adjustments are to be made once the study begins. Stakeholders should also discuss supporting and adopting adaptive designs thereby allow decision-making after planned interim analysis of *intervention* data, to either stop the trial for futility or to continue the trial with adjusted design parameters, such as sample size.^62, 63^ Perhaps just as important in the context of vector control, although RCTs are rated as high-quality evidence,^64^ considerations to RCT alternatives are prudent in the trial planning phase as these may offer assurances of adequate data rigor while balancing cost and time constraints of traditional RCTs.^65, 66^ Alternative study designs include large observational studies for detecting population-level effects using analytical cross-sectional studies, or operational program-based evidence; the latter perhaps especially for interventions containing an existing registered chemical active ingredient (i.e., meets human safety thresholds) and where the intervention is implemented in pilot trials and impact monitored through case reports, as compared to a contemporaneous control group.^66^

Albeit the study was not powered for entomology, the inclusion of entomological endpoints in the Sumba RCT was, in part, to help understand and validate the intervention’s mode of action for the VCAG claim of a health impact through a reduction in human-vector contact. Spatial repellent chemicals may cause initial knockdown and mortality by exposure to toxic doses at close range to the active ingredient, or a delayed kill through behavioral avoidance response (i.e., blood-feeding inhibition), through exposure to sub-lethal doses at distances further away from the stimulus source. Therefore, entomological measurements for detecting reductions in human-landing density in clusters with active intervention, and a possible change in anopheline age structure (parity rate) indicating reduced daily survival of mosquitoes in clusters with intervention, was built into this study. Additionally, a reduction in sporozoite infection rates, because of lower blood feeding success, and/or an inability to survive the required time interval (parasite incubation period) to become infective (transmissible) from a vector to a host, represent other endpoint measures of repellency impact. A causal relationship with one, several or all, of these endpoints would allow future trials evaluating non-inferiority of a ‘second-in-class’ spatial repellent to integrate entomological measures only to predict PE and provide assurances of meeting minimum thresholds of acceptance for public health use. The value arguably, is a potential reduction in cost and time which subsequently could further incentivize industry R&D with the goal of increasing varying types of quality, efficacious spatial repellent products available for global implementation.

Sporozoite positivity rates and EIR estimates from the current trial were low, with a total of 42 of 11,650 (0.36%) anophelines tested during baseline (10mo) and 19 of 17,971 (0.11%) found infected during intervention (24mo). These findings are not unusual or unexpected in low to moderate malaria transmission settings.^35^ As example, the previous Sumba study (using metofluthrin coils) reported just 15 out of 1,825 (0.82%) HLC anopheline samples sporozoite infected.^19^ The challenges for assessing sporozoite infectivity and using EIR as a measure of intervention effect in such settings should be factored into study pre-planning to carefully balance cost of sample processing with potential useful information gained.

These study findings report a 16.4% numerical reduction in indoor-landing mosquitoes exposed to active treatment compared to placebo-control. Just as important was an observed numerical reduction (11%) on outdoor-landing collections on the exposed verandah of sentinel houses, as Sumba residents often sleep on the verandah without the protection of a bed net. It is noted, however, that both indoor and outdoor HLC outcomes showed wide CIs that overlap zero, making results inconclusive; nevertheless the intervention showed impact against malaria in these clusters. A previous proof-of-concept study on Sumba that evaluated a metofluthrin-active coil resulted in a significant 32% reduction in *An. sundaicus* indoor landing rates as compared to a blank-coil control;^19^ The minimal reduction in the human landing rate in the current RCT may be the result of species-specific effects by active ingredient (i.e., metofluthrin vs. transfluthrin) but differences in HLC outcomes of the two studies on Sumba should not be interpreted as the transfluthrin intervention being less efficacious than a coil. The greater complexity of anopheline diversity may likely have contributed to the inconclusive HLC outcomes based on aggregated anopheline data. It is expected similar results may occur in future trials that are conducted in settings with diverse vector populations and/or as cluster size increases thereby increasing the probability of greater habitat diversity.

Specifically, the previous Sumba study was smaller in scale and *An. sundaicus* was the overwhelmingly predominate human-feeding anopheline collected; whereas in the current study this species was relatively uncommon during the intervention trial. The range of ecologies (coastal plains to upland forested hills) varied greatly across the 12 clusters used for entomology collections in this trial compared to that of the earlier Sumba study where collections were confined to only 4 adjoining clusters having similar coastal environments. This spatial variability in ecology contributed to a broader range of species diversity. We report 19 species/group species identified), each species with different bionomic and behavioral characteristics (e.g., biting habits, host preferences). Perhaps most important, the efficacy of an intervention is related to epidemiological outcomes (reduced risk of infection); HLC outcomes reported from the current study were inconclusive, but nevertheless showed impact against malaria.

Insecticide susceptibility monitoring was integrated into the study to characterize the wild-type anopheline populations to estimate PE against resistant vector populations, if resistance was expressed, and monitor changes in susceptibility due to continuous exposure to transfluthrin. Our data indicates that continual use of transfluthrin as a spatial repellent during a consecutive 24-mo period did not result in a change in phenotypic response to permethrin. Although we are confident in the findings presented, there were limitations in sampling from multiple sites and handling numerous species during pre-, within, and post-intervention periods. This may have compromised the study’s ability to obtain a more robust and definitive profile of susceptibility background and detection of any shift in phenotypic (and molecular) frequencies.

The third ‘lesson-learned’ from the current study relates to AE and SAE monitoring. Reports of intervention safety in this study should take into account possible limitations of the data collection. The DSMB was provided with clinic attendance data from January 2015 to December 2018, for upper respiratory infection (URI), pneumonia, and malaria, as classified by month, health center, and village within health center coverage. Although not gathered as part of the study, the DSMB was keenly interested in these data because of the surprisingly low number of illnesses/deaths reported from the study population during the 24-month intervention period. For some periods, particularly for malaria infection, the case data were missing or the health center totals were not available at the village level. Of the 13 villages represented in the dataset, from which study clusters were formed, all but four comprised clusters (or parts of clusters) from both study intervention arms (i.e., allocated active or placebo); therefore, it was not possible to infer the allocation status of the corresponding cases. For this reason, the DSMB was unable to make a reliable assessment of any possible association between active intervention and clinic attendance for those reported health conditions. The respiratory illnesses in the clinic data are more reflective of the magnitude of numbers of cases one would expect from this population; however, the DSMB was again unable to parse them into test and control clusters due to reasons stated above, and therefore the data was of little use for monitoring purposes. The number of SAEs reported, a total of six, is well below the expected in a study population of this size. The WHO estimates the crude death rate in Indonesia to be approximately 6.2 per thousand per year.^67^ Based on this, for a population of 1,296 children enrolled (protocol sample size), the probability of having zero deaths in a year is less than one in 1,000. Although open commemoration of deaths is commonly practiced in the study area, it seems more likely deaths were not completely reported/recorded as SAEs.

Overall, the point to apply for future spatial repellent trials (and perhaps other trials of new vector control interventions) is to improve the mechanism for capturing and communicating AEs and SAEs before the initiation of the study to better ensure reliable reporting and regular monitoring for unusual numbers of complaints. If these studies are regarded as a clinical trial with a placebo arm that requires comparative AE and SAE data, the scale could cause failure in safety assessment simply on the mass of data, however imperfectly collected. Any safety signal detected would most likely be due to bias in reporting and/or collection bias and unrelated with the investigational intervention. Trials evaluating a spatial repellent intervention with a registered active ingredient (i.e., a chemical meeting regulatory approvals for acceptable levels of human risk) such as transfluthrin, could focus on a small number of complaints that might be expected due to inhalation (i.e., volatilization) and develop a monitoring scheme that collects consistent data across the study population. Clinic-based complaints of respiratory illness such as pneumonia and asthma might be possible, but the variability in their method of collection (clinic or survey) will likely be highly dependent on study site infrastructure.

In conclusion, while more evidence will be required to determine whether spatial repellents can serve as a viable malaria control intervention, both the primary and secondary results of this Sumba Island trial have generated valuable data and observations that can contribute to the overall assessment and improvement of testing protocols of a spatial repellent intervention class. The VCAG has recommended that data from at least one additional trial evaluating the spatial repellent be generated,^68^ and once available, the panel will be able to assess the available evidence for judging public health value. If the crude estimate of PE shown here, near 30%, is replicated in future statistically robust cluster-randomized trials, the intervention prototype evaluated in this RCT would approximate the benefit associated with LLINs.^69^ Perhaps just as important, the number of cases averted indicates 361 expected cases per 1000 persons per year (0.361*1000*1) without the spatial repellent and 152 less cases expected when using the active intervention ((0.361-0.209)*1000*1). The cost-savings of averting these 152 less cases per 1000 person-years is an important consideration for health-systems strengthening. These results have encouraged further and substantial investment to validate spatial repellent efficacy through larger RCTs, including an investigation of possible vector diversionary effects on human health (i.e., greater than expected malaria incidence in households near intervention not receiving active product), and evaluation of the optimal delivery systems for humanitarian assistance use-case scenarios.^70^

## Data Availability

Analytical datasets available upon request to corresponding author.

## Acknowledgments

We extend sincere appreciation to DSMB members: Dennis Shanks, Chris Drakeley, and Neal Alexander for assurances on data integrity and safety assessments. We also express our gratitude to fhi360 partners Kathy Bowerman, Ahmed Goolam and Zen Hafy for trial oversight and monitoring of protocol compliance. We thank the WHO VCAG for their comments from recurrent project data assessments. We are grateful for the tireless efforts of Wirda Damanik, study administrator at the Eijkman Institute for Molecular Biology in Jakarta, and Marianne Kent, Program Coordinator at the University of Notre Dame, for their expert administrative, logistical, and financial management support of this endeavor. We thank Julie Neidbalski, Katie Cybulski, Diane Lovin, Joanne Cunningham, Jenna Davidson, and Allison Hendershot at the University of Notre Dame for their contribution to human blood and mosquito sample quality control processing as well as David Pettifor of the University of Notre Dame Center for Research Computing for assisting with central database development and data verification. We gratefully thank the residents in the study area for their time and patience participating in this trial. Special gratitude is extended to the Southwest and West Sumba Districts Health Department for their kind support, the parasitology and entomology teams, local field workers, data entry clerks, and the numerous local volunteers for their long-serving dedication to quality of data and meeting the project objectives.

## Disclaimer

The contents are the responsibility of the authors.

